# Quantum Machine Learning and Data Re-Uploading: Evaluation on Benchmark and Laboratory Medicine Datasets

**DOI:** 10.1101/2025.05.14.25327605

**Authors:** Thomas JS Durant, Seung Joo Lee, Sarah Dudgeon, Elizabeth Knight, Brent Nelson, H. Patrick Young, Lucila Ohno-Machado, Wade L. Schulz

**Affiliations:** Department of Laboratory Medicine, Yale School of Medicine, New Haven, CT; Biomedical Informatics and Data Science, Yale School of Medicine, New Haven, CT; Yale School of Medicine, Yale University, New Haven, CT; Computational Biology and Bioinformatics, Yale University, New Haven, CT; Newport Healthcare, Minneapolis, MN; Department of Psychiatry, University of Minnesota, Minneapolis, MN

## Abstract

**Background:** Quantum machine learning is an emerging field that may offer unique advantages over classical machine learning but has not been extensively studied with real-world healthcare data of practical size. This study evaluates the performance of a recently proposed quantum machine learning algorithm—quantum circuits with data re-uploading—compared to classical machine learning and other common quantum algorithms, using both benchmarking data and real-world laboratory medicine data.

**Methods:** Four datasets containing between 2 and 30 features were selected for evaluation. The quantum re-uploading algorithm was compared against variational quantum classifiers, quantum neural networks and four, commonly used classical machine learning algorithms. Initial, baseline comparisons of classification performance (F1 score) were conducted using all algorithms across the four datasets.

Configuration parameters were then optimized for the quantum data re-uploading algorithm using a previously published dataset of plasma amino acid profiles to determine the impact of optimization on classification performance, followed by a final comparison against classical ML algorithms.

**Results:** Baseline comparisons of quantum re-uploading showed superior classification performance on lower-dimensional datasets compared to quantum and linear ML algorithms. However, performance declined significantly as the number of input features increased. While optimization improved classification performance, marked variability was observed. Following optimization, the quantum re-uploading algorithm again performed comparably to linear algorithms but had lower classification performance on the plasma amino acid dataset compared to non-linear classical algorithms.

**Conclusion:** This study suggests that quantum data-reuploading algorithms can achieve classification performance comparable to some classical methods on lower-dimensional datasets, presenting opportunities for early research applications in laboratory medicine. However, while optimizing configuration parameters can improve classification performance, further advancements in quantum hardware and algorithm design will likely be necessary for quantum machine learning to become practically viable in laboratory medicine, and biomedical research more broadly.

## INTRODUCTION

In the era of personalized medicine, there is growing interest in using data analytics to deliver more tailored diagnoses and treatments. Today, artificial intelligence (AI) and machine learning (ML) play important roles in these efforts and have been widely implemented in various clinical specialties, including laboratory medicine and its subspecialties (1–3). ML has been successfully applied to diverse tasks in the clinical laboratory, such as image classification, automated quality assurance, and disease risk prediction (2,4). However, current ML algorithms face challenges in maintaining interpretability, generalizing effectively from smaller datasets, and ensuring equitable predictive performance across demographic subgroups (5–7). Consequently, there is an ongoing need to evaluate novel ML algorithms and explore solutions that may enhance the capabilities of ML applications in healthcare and laboratory medicine.

Quantum computing is an emerging field that offers theoretical advantages over classical computing (8,9). By leveraging basic principles of quantum mechanics, quantum computers (QCs) can represent multiple computational states simultaneously (9). This capacity allows QCs to theoretically perform calculations more efficiently than classical computers (10,11). Over the last decade, access to QCs has increased with many available through public cloud platforms with vendors offering ongoing improvements in QPU size and error mitigation strategies (8). In addition, the development of open- source quantum software development kits (QSDKs) and quantum processing unit (QPU) simulators have allowed for increased adoption across many scientific domains. As a result, there has been a growing number of publications describing novel quantum algorithms and investigating the potential use of QCs for practical applications, using both QPU simulators and actual QPUs (8).

Quantum machine learning (QML) is a subfield of quantum computing that both adapts classical ML methods to quantum frameworks and introduces quantum-native algorithms that do not have direct classical counterparts (8,12). Recent research has investigated the utility of quantum computing for common ML tasks such as classification, regression, clustering, and feature selection (13–15). While some of these studies report promising classification performance on low-dimensional datasets, few consider higher-dimensional, non-synthetic datasets commonly encountered in real-world applications. This is understandable, as increasing the number of qubits in a quantum circuit makes the system more susceptible to noise and decoherence, which can introduce errors during computation (9,13,16). This limitation can be more impactful on some QML algorithms than others, such as quantum neural networks (QNNs), which require larger quantum circuits (i.e., more qubits) for datasets with more input features (8,17). Quantum simulators are also insufficient in these situations because they require a large amount of memory to simulate QPUs on classical hardware. To address these limitations, novel approaches to quantum algorithm and circuit design have recently been proposed.

In 2020, Pérez-Salinas et al. described a novel algorithm for quantum ML classification tasks that requires minimal quantum resources and smaller quantum circuits relative to other QML algorithms, such as quantum neural networks (18). They showed that a single qubit, combined with data re-uploading, can function as a universal classifier and may enable the approximation of continuous functions (18). With fewer qubits required, this approach offers a valuable opportunity to evaluate QML classification performance on practical, higher dimensional datasets while still leveraging quantum properties that may offer advantages over classical ML. Numerous publications have since explored QML and data re-uploading but with a primary focus on mathematical theory, proofs related to the universal approximation theorem, applications to lower-dimensional synthetic datasets, or higher-dimensional datasets with the application of dimensionality reduction techniques (19–22). While these works provide valuable insights into potential uses for binary and multi-class classification problems, there is still a need to investigate how this method performs using data which are non-synthetic, clinical datasets.

In this study, we assessed the performance of quantum circuits with data re-uploading (QC- REUP) compared to state-of-the-art classical and quantum machine learning algorithms on benchmark datasets. We also systematically examined different configurations of QC-REUP, evaluating the effects of circuit size, qubit entanglement, and data re-uploading strategies on classification performance. To assess the short-term potential of QML for real-world healthcare data, we applied these methods to a previously published, clinical laboratory dataset from Wilkes et al., which includes 28 input features from plasma amino acid (PAA) profiles obtained during routine clinical testing for inherited metabolic diseases (23). The evaluation of the PAA dataset represents a practical biomedical QML application that has been benchmarked using classical ML methods (23,24).

## METHODS

### Technology

All experiments used Python (version 3.10) within version-controlled, virtualized Miniconda environments. Quantum-based algorithms were implemented using the Qiskit SDK (version 1.0.2; IBM, Armonk, NY). A grid search of all configuration parameters was conducted and logged using the Azure Machine Learning/mlflow library (version 2.12.2). All analyses were run on Azure serverless virtual machines equipped with traditional CPUs (48 cores, 96 GB RAM, 384 GB disk). All QML algorithms in this study were executed using quantum simulators on the same serverless virtual machines. Classical ML algorithms used scikit-learn (version 1.4.2), xgboost (version 2.0.3), and tensorflow (version 2.16.1).

Feature scaling methods were implemented for continuous variables using the sci-kit learn package (version 1.4.2). T-tests were performed with the python package SciPy (version 1.13.0).

### Dataset Selection

We evaluated the performance of classical and quantum machine learning algorithms using four datasets (**Figure 1**). These datasets were selected to represent both low- and high-dimensional input feature spaces and are well-documented in the literature regarding classical ML performance. Three of the datasets are commonly used for ML benchmarking: the (1) Circle, (2) Iris, and (3) Wisconsin Diagnostic Breast Cancer (WDBC) datasets (25,26).

**Figure 1:**
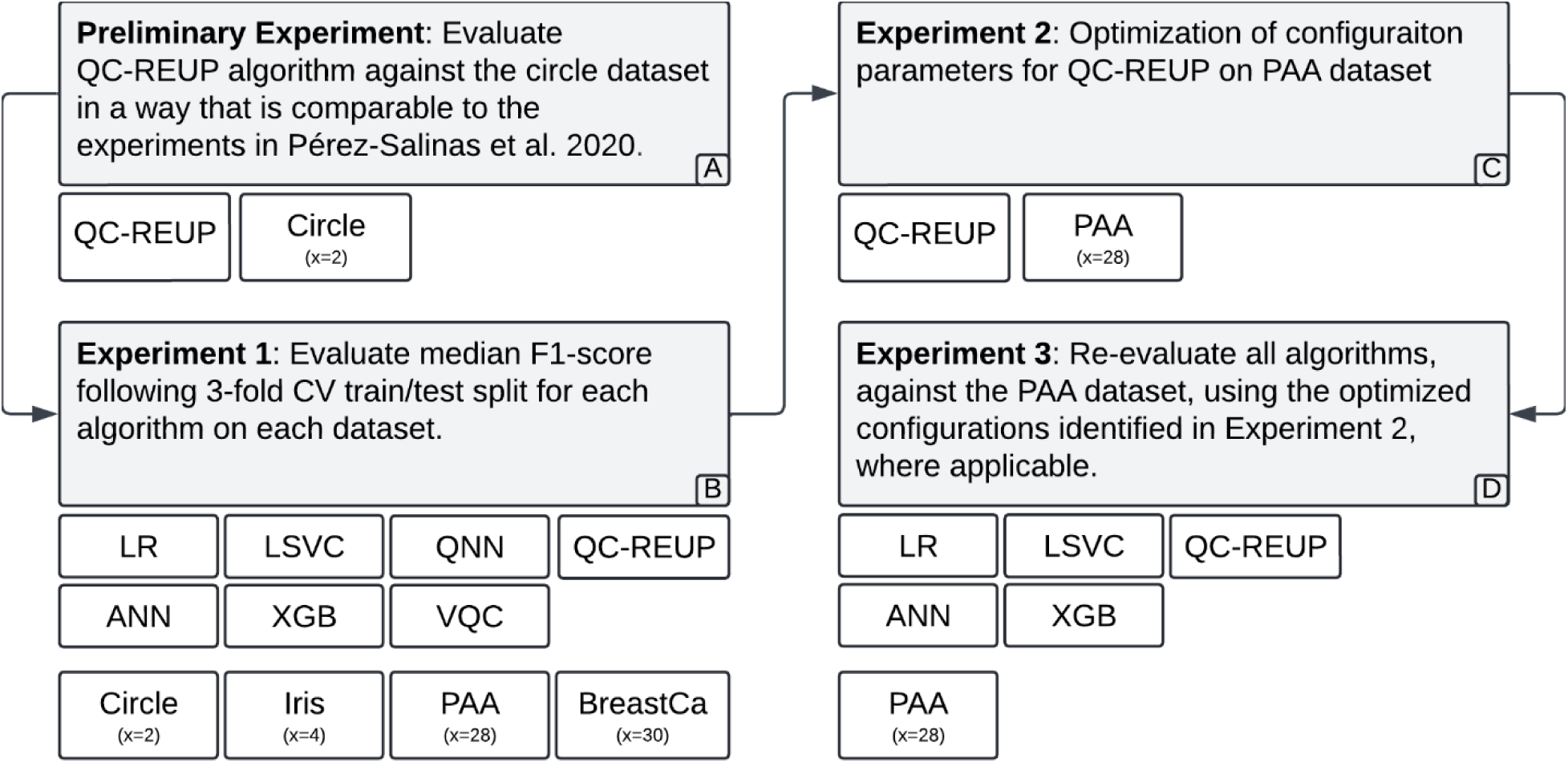
Diagram of overall study design and experiment flow.

The Circle dataset is a synthetic, two-dimensional dataset with a binary classification where the classes are separated by a single circular boundary (**Supplemental Figure 1**). The Iris dataset is widely used in ML and contains 150 samples from three species of iris flowers, with four features (sepal length, sepal width, petal length, and petal width). For the Iris dataset, two of the three mutually exclusive classes were combined into one class to convert the data into a binary classification problem. The WDBC dataset consists of 569 instances with 30 features derived from digitized images of fine needle aspirate samples of breast masses with a binary classification of benign or malignant. The fourth dataset, the PAA dataset, was obtained from routine clinical chemistry testing as described by Wilkes et al. It contains 2,084 anonymized PAA profiles, each with 28 features, and is enriched for rare disease cases (23). Each record is labeled as either normal or abnormal.

### Algorithm Selection

For comparison against QC-REUP, qe selected four classical ML algorithms for this study, with the intent to include those which are capable of learning linear and non-linear decision boundaries. Specifically, we evaluated the performance of logistic regression, linear support vector classifier (LSVC), extreme gradient boosting (XGB), and an artificial neural network (ANN). For QML algorithms, we implemented a variational quantum classifier (VQC) and a quantum neural network (QNN).

In this study, a custom implementation of the QC-REUP algorithm, as outlined by Pérez-Salinas et al., was developed by the authors, using the Qiskit library and implemented in Python. The QC-REUP algorithm followed the architecture described by Pérez-Salinas et al. and used a single qubit and two data reupload layers for the initial baseline comparison between classical and quantum ML algorithms (**Figure 2**) (18). Configurations for the QNN algorithm were chosen based on configuration parameter optimization performed in a prior study by the current authors. The ANN algorithm architecture was a fully connected, feed-forward neural network with an input layer, followed by a single hidden layer containing 64 neurons activated by ReLU. The output layer was a single neuron with a sigmoid activation function, and the network was compiled for binary classification using the Adam optimizer, binary cross entropy loss, and accuracy as the performance metric. All other algorithms were implemented using default settings unless otherwise described.

**Figure 2:**
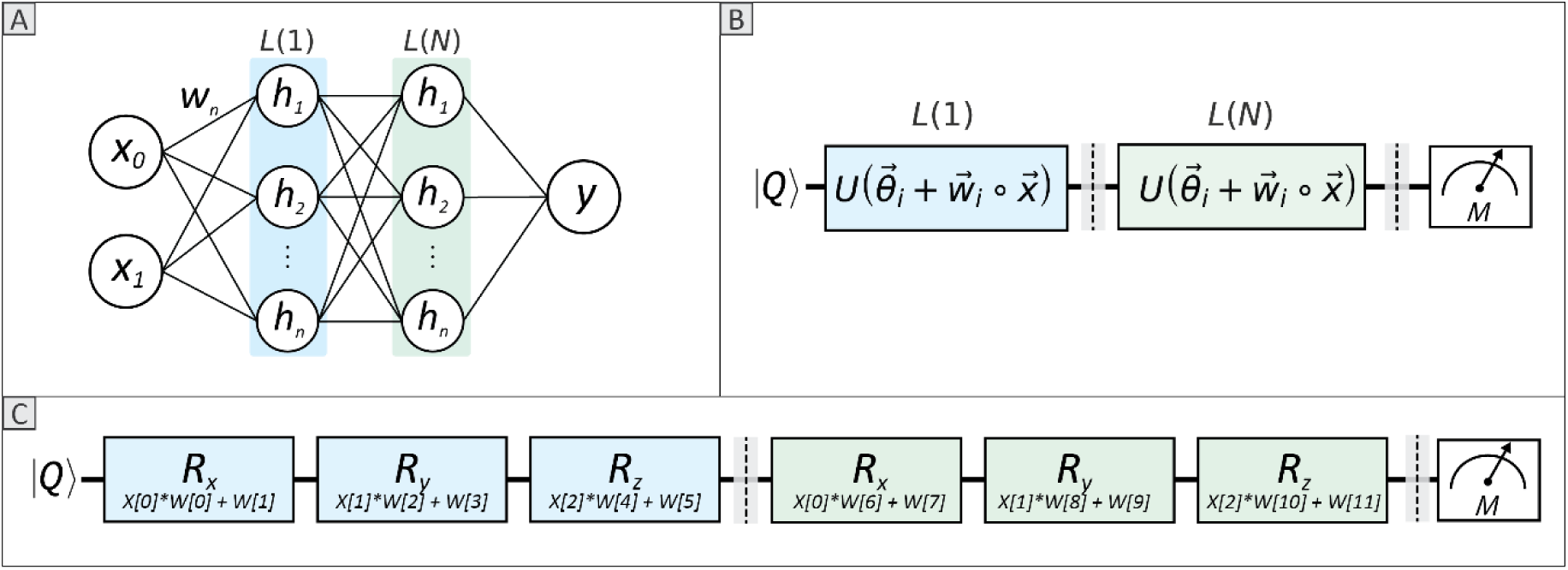
**(A)** Diagram of a fully-connected, feedforward artificial neural network where input features (*x*) are received as weighted inputs (*w*) into neurons (*h*) arranged in layers (*L*), where *L*_1_ is shaded blue, and Ln is shaded green. Each neuron (box) applies a transformation (e.g., weighted sum and activation function) and passes the result to the next layer. **(B)** Quantum circuit diagram for data reuploading with a single qubit register (*Q*). Classical input features are encoded via parameterized unitary transformations (*U(Ɵ_*i*_ + w_i_* ∘ *x)*), arranged in layers (*L*), and measured (*M*). **(C)** Explicit encoding of features (*x_i_*) into quantum gates (R_x_, R_y_, and R_z_), applied to qubit (*Q*), with their respective rotational planes (e.g., X, Y, or Z-plane). Input (*X*) and weight (*W*) arrays with the associated indices, indicate where features and weights are applied. Dashed lines mark boundaries of ‘re-upload layers’. Taken together, this figure illustrates a quantum circuit with a single qubit and two data reupload layers, as it compares to a conceptually analogous ANN architecture.

### Experimental Design: Classical and Quantum Machine Learning Comparison

The overall study design and experiment flow can be visualized in **Figure 1**. Unless otherwise noted, all experiments used a 70:30 train-test split and 3-fold cross-validation to assess performance variability on the test dataset. All experiments use the F1 score, or additional metrics if specified, and were calculated from predictions on the test dataset.

To verify the correctness of the author’s QC-REUP implementation, the algorithm was first tested on the Circle dataset following a similar approach to that described by Pérez-Salinas et al. (**Figure 1A**).

Classical ML and QML algorithms were then evaluated on each dataset and compared by F1 score (**Figure 1B**). Following this baseline comparison, optimization of configuration parameters was performed for the QC-REUP algorithm using the PAA dataset (**Figure 1C**). Once the optimal configurations were determined, the QC-REUP was again compared to classical ML algorithms, this time only using the PAA dataset (**Figure 1D**). To provide a more balanced comparison with the classical ANN algorithm, layer architecture was adjusted to approximately match the number of trainable parameters in the QC-REUP algorithm. A paired one-tailed t-test was conducted to compare the F1 score of the QC- REUP algorithm against the classical ML algorithms, across five-fold CV trials to assess whether classical algorithms significantly outperformed QC-REUP using optimized configurations. Statistical significance by t-test was evaluated at 0.0125 after Bonferroni correction.

### Quantum Circuit Data Re-Uploading Configuration Parameter Optimization

In this study, configuration parameters refer to settings established before training that can affect the model’s performance. These parameters include those related to dataset preparation (e.g., feature scaling), quantum circuit design, and the method by which data is uploaded to the quantum circuit (**Table 1**). Data re-uploading methods include symmetrical and asymmetrical approaches and are applicable only when there is more than one qubit. These methods determine how input features are distributed across multiple qubits. For instance, in a quantum circuit with more than one qubit, a symmetrical re-upload approach applies each input feature (*xi*) to all qubits. In contrast, an asymmetrical approach sequentially uploads input features to qubit 1 and then to qubit 2, ensuring equal distribution across the available qubits.

**Table 1:** Options for quantum circuit data re-upload configuration parameter grid-search.

| Configuration Parameter | Options |
| --- | --- |
| *Downsample Negative Class* | True, False |
| *Feature Scaler* | standard, min0max1 |
| *Number of Qubits* | 1, 2, 4 |
| *Number of Re-Upload Layers* | 2, 4, 6, 8 |
| *Quantum Entanglement* | True, False |
| *Quantum Re-upload Method* | Symmetrical, Asymmetrical |
| *Quantum Noise* | True, False |

To evaluate the impact of each configuration parameter on QC-REUP classification performance by F1 score, we analyzed Shapley Additive Explanations (SHAP) values and feature importance by random forest regression. In addition, we examined the frequency of each parameter value’s occurrence within the top 10% of grid-search runs, as ranked by F1 score.

### Data and Code Availability

The Circle dataset was generated as described in the previous section and is available as part of the project’s code. The Iris and WDBC benchmark ML datasets are publicly available from various sources (27). PAA data are available from the original publication (23). The code was independently written and reviewed by two authors.

## RESULTS

### Initial Classical and Quantum Machine Learning Comparison

The QC-REUP algorithm we implemented demonstrated comparable performance to the originally described implementation on the circle dataset (**Supplemental Table 1**). For the initial baseline comparison between classical ML and QML models (**Figure 1B**), the QC-REUP model demonstrated comparable performance to the classical ML algorithms on lower-dimensional datasets, with QC-REUP demonstrating a median F1 scores of 0.90 (95% CI: 0.88-0.91) and 0.86 (95% CI: 0.83-0.96) on the Circle and Iris datasets, respectively. However, the QC-REUP model demonstrated a significant decline in classification performance with higher-dimensional input feature spaces with median F1 scores of 0.40 (95% CI: 0.36 - 0.41) and 0.44 (95% CI: 0.41 - 0.45) on the PAA and WDBC datasets, respectively. Review of the QC-REUP training loss values demonstrated a failure to minimize on the PAA dataset (**Supplemental Figure 2**). Some algorithms, such as the QNN and VQC, failed to converge or exceeded computational resources and therefore did not finish training on higher-dimensional datasets (**Table 2**).

**Table 2:** Median F1 score across 3-fold cross validation with associated 95% confidence interval. Number of input features are indicating in parentheses next to dataset.

| Algorithm | Dataset |  |  |  |
| --- | --- | --- | --- | --- |
|  | Circle (2) | Iris (4) | PAA (28) | WDBC (30) |
| Linear Regression | 0.51 (0.51-0.54) | 0.68 (0.60-0.74) | 0.62 (0.60-0.62) | 0.91 (0.91-0.96) |
| Linear SVC | 0.51 (0.51-0.54) | 0.72 (0.58-0.75) | 0.45 (0.45-0.62) | 0.87 (0.81-0.91) |
| Neural Network | 0.98 (0.98-0.99) | 0.87 (0.83-0.93) | 0.70 (0.70-0.74) | 0.92 (0.91-0.96) |
| XGBoost | 0.98 (0.97-0.98) | 0.90 (0.89-0.96) | 0.85 (0.84-0.85) | 0.96 (0.92-0.98) |
| QC-REUP | 0.90 (0.88-0.91) | 0.86 (0.83-0.96) | **0.40 (0.36-0.41)** | 0.44 (0.41-0.45) |
| QNN | 0.85 (0.85-0.87) | 0.63 (0.37-0.71) | DNF | DNF |
| VQC | 0.86 (0.86-0.87) | 0.48 (0.48-0.71) | DNF | DNF |

Abbreviations: DNF = Did not finish; SVC = Support Vector Classifier; XGBoost = Extreme Gradient Boosting; QNN = Quantum Neural Network; VQC =

### Quantum Circuit Data Re-Uploading Configuration Parameter Optimization

Using the pre-defined configuration parameter search space (**Table 1**), approximately 200 unique runs were conducted with the QC-REUP algorithm on the PAA dataset (**Figure 1C**). Runs using a noisy quantum simulator were excluded from the final analysis due to these requiring excessive computational resources and computation time (>48 hours). The median F1 scores across all runs ranged from 0.13 to 0.64, with a median of 0.40 (95% CI: 0.19-0.61) (**Figure 3A**). SHAP values indicated that downsampling the negative class, feature scaling, and the number of qubits were the most influential factors on F1 score (**Figure 3B**). This was also corroborated by the feature importance analysis by random forest regression (**Supplemental Table 3**). The frequency of parameters in the top 10th percentile of F1 scores aligned with the SHAP analysis; however, showed a slight advantage for the data scaling (range 0-1) (**Figure 3C**). In addition, increasing the number qubits generally had a negative impact on F1 scores, while increasing the number of layers had mixed impact.

**Figure 3:**
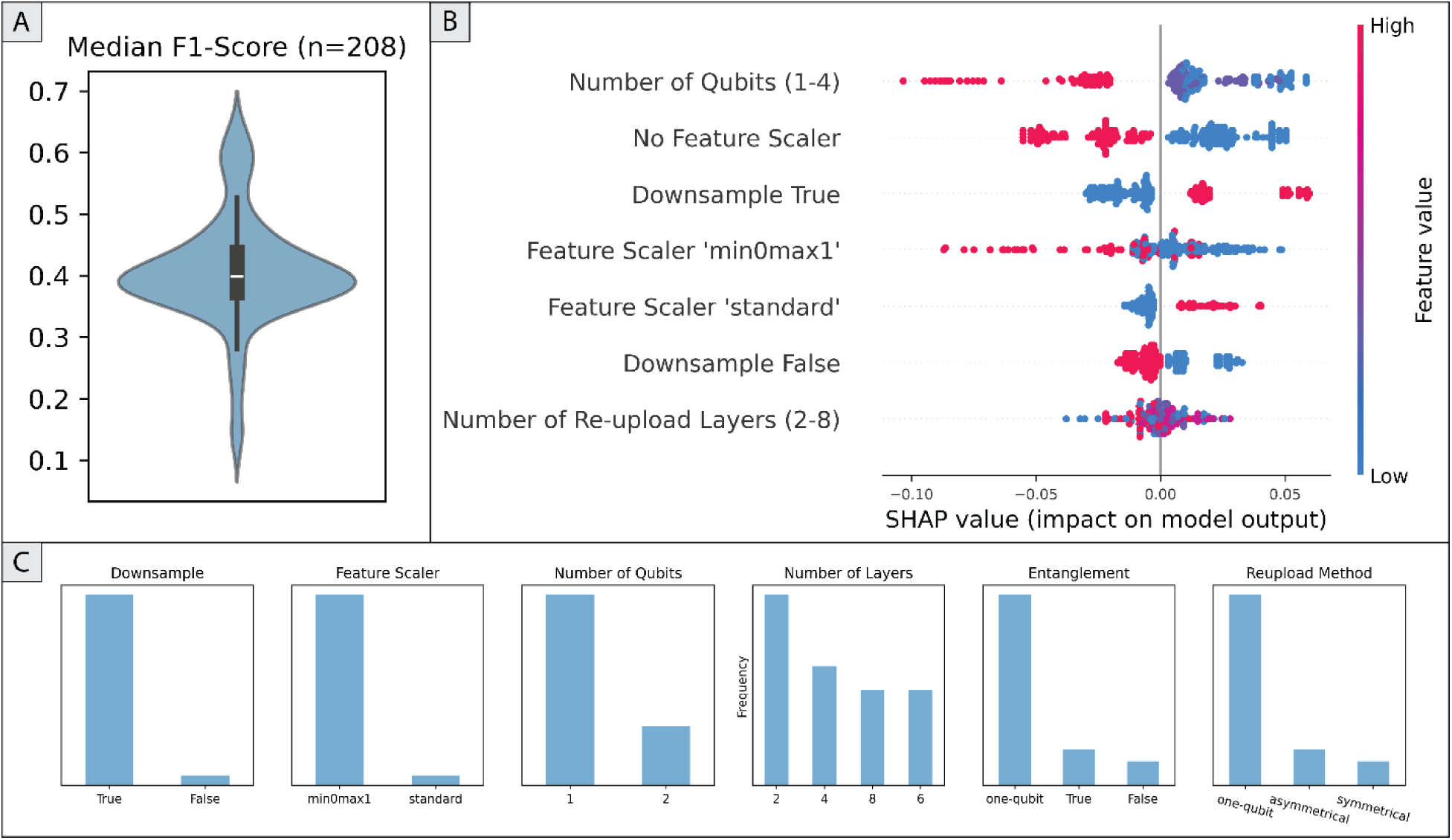
(A) Violin plot of median F1 score for QC-REUP algorithm on PAA dataset for all trials that completed successfully. (B) SHAP Summary plot for quantum circuit data re-uploading configuration parameter optimization. Binary features are one-hot encoded, and features with minimal or no contribution are omitted. (C) Frequency of configuration parameters among the top 10% of sweep trials based on median F1 score.

### Optimized Quantum Circuit Data Re-Uploading Comparison on PAA Dataset

The comparison of the classical ML algorithms to the QC-REUP, using the PAA dataset, used the following configuration parameters based on our optimization results: negative class downsampling, data scaling (range 0-1), one qubit, and six data-reupload layers (**Supplemental Figure 3**). Examination of the QC- REUP train loss values demonstrated an appreciable decrease over training iterations, indicating optimization convergence (**Figure 4**). The QC-REUP algorithm demonstrated a median classification accuracy of 0.74 (95% CI: 0.64-0.79) and an improved median F1-score of 0.61 (95% CI: 0.48-0.64). The difference in classification performance between QC-REUP and logistic regression or LSVC was not statistically significant. However, classification performance of ANN and XGB demonstrated significantly higher classification performance based on F1 score (**Table 3**).

**Figure 4:**
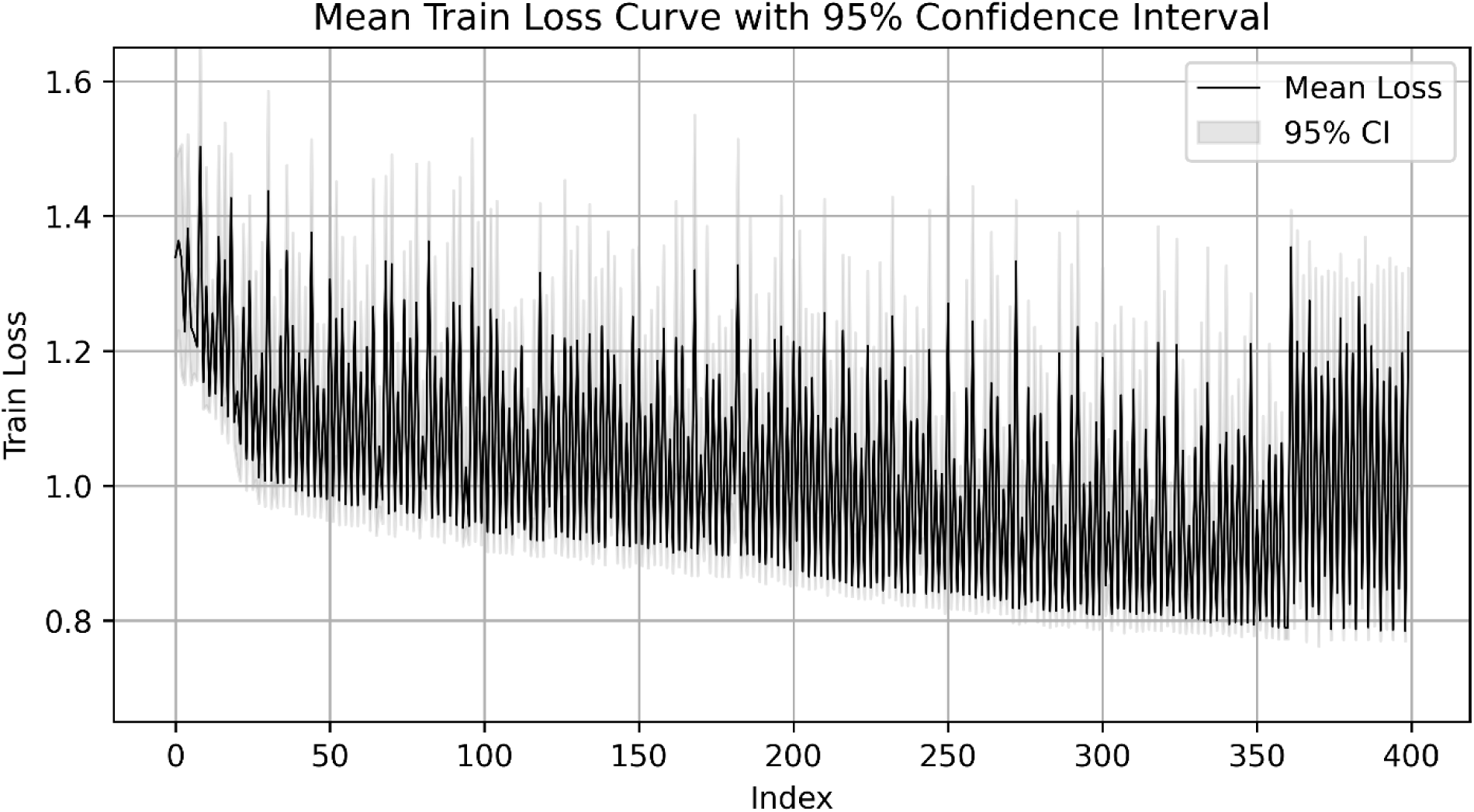
Mean training loss curve with 95% confidence interval for the quantum circuit data re- uploading algorithm on the PAA dataset.

**Table 3:** Median and 95^th^ percentile for specified performance metrics on the plasma amino acid dataset across 5-fold cross validation. ¥ denotes p-value < 0.0125 as compared to QC-REUP median F1 score classification performance.

| Algorithm | Accuracy | Recall | Precision | F1 Score |
| --- | --- | --- | --- | --- |
| *Linear Regression* | 0.77 (0.75-0.79) | 0.65 (0.59-0.67) | 0.61 (0.59-0.67) | 0.64 (0.59-0.66) |
| *Linear SVC* | 0.79 (0.78-0.81) | 0.65 (0.60-0.69) | 0.66 (0.64-0.70) | 0.65 (0.63-0.70) |
| *Neural Network* | 0.86 (0.85-0.90) | 0.70 (0.66-0.81) | 0.81 (0.80-0.86) | 0.75 (0.73-0.83)¥ |
| *XGBoost* | 0.91 (0.90-0.93) | 0.90 (0.88-0.93) | 0.83 (0.78-0.87) | 0.86 (0.85-0.89)¥ |
| *QC-REUP* | 0.74 (0.64-0.79) | 0.60 (0.53-0.71) | 0.58 (0.43-0.67) | **0.61 (0.48-0.64)** |

Abbreviations: SVC = Support Vector Classifier; XGBoost = Extreme Gradient Boosting; QC-REUP = Quantum Circuit Re-Upload

## DISCUSSION

In this study, we evaluated QC-REUP against other QML and classical ML algorithms on commonly used benchmark datasets, including a previously published dataset derived from routine clinical care (23). Our results indicate that QML algorithms, particularly QC-REUP, can achieve classification performance that is comparable to classical ML counterparts with lower-dimensional datasets, mirroring results from prior studies (18,28). More generally, these findings are aligned with previous reports that QML algorithms are capable of approximating non-linear functions encountered with lower complexity datasets (18,29).

However, our data also show that classification performance of QML algorithms, including the data reuploading strategy, can decline when applied to more complex and higher-dimensional feature spaces, such as those often generated in healthcare and biomedicine.

In our initial evaluation (**Figure 1B**), the QNN and VQC algorithms demonstrated a decline in classification performance between the circle and the iris datasets, as the number of input features increased from two to four, respectively. Further, these algorithms failed to train successfully on both the PAA and WDBC datasets due to convergence issues or excessive computational time. The inability to complete training on the larger datasets for QNN and VQC was somewhat expected due to the one-to-one mapping of input feature to qubits for the architectures used in this study, which leads to wider quantum circuits (i.e., more qubits) and requires substantially more computational resources for quantum simulation (8). In contrast, the QC-REUP algorithm achieved higher F1 scores than other quantum algorithms on the lower-dimensional Circle and Iris datasets, maintaining relatively consistent performance between both. However, its classification performance declined significantly on the higher-dimensional WDBC and PAA datasets. The cause of this decline as the input feature space increased from 4 to 30 remains unclear, but it may indicate a fundamental limitation of the re-uploading approach in learning non-linear decision boundaries in higher-dimensional spaces.

Optimization of configuration parameters for the QC-REUP algorithm on the PAA dataset (**Figure 1C**) led to some improvement in classification performance but also highlighted challenges in tuning QML algorithms. The wide variation in F1 scores (0.12 to 0.64) underscores the algorithm’s sensitivity to configuration parameter choices. Feature scaling and downsampling the negative class during data preprocessing were among the most influential factors affecting classification performance. These findings suggest that the data re-uploading architecture is sensitive to input feature range and class imbalance. Additionally, we observed that increasing the number of qubits generally decreased classification performance, while increasing the number of layers had a more variable effect on F1 scores. Under an ideal QPU simulator—where quantum noise and decoherence do not contribute to information loss—this was an unexpected result, as more trainable parameters would typically be expected to improve model performance.

In classical machine learning, classification performance can be improved by tuning configuration parameters during the training process (30). While this can lead to notable performance improvement on test sets, the variation reported in most classical ML literature is generally not as pronounced as what we observed in our QML experiments (20,31,32). Of note, a similar degree of variation and sensitivity in classification performance to parameter choices was observed in our previous work with QNNs. Although a direct comparison with classical ML parameter optimization was outside the scope of this study, it should be acknowledged that the exact degree of performance variability likely depends on the specific dataset, model, and experimental setup. Nevertheless, these observations underscore the importance of incorporating similar evaluation and optimization protocols in future QML research.

Despite the challenges encountered, optimization of configuration parameters for the QC-REUP algorithm was able to significantly improve classification performance on the PAA dataset, with an increase in median F1 score from 0.40 (95% CI: 0.36 - 0.41) to 0.61 (95% CI: 0.48-0.64). When considering classification performance relative to classical ML algorithms (**Figure 1D**), the median F1 score of the optimized QC-REUP algorithm was not significantly different as compared to linear classical ML algorithms (**Table 3**). For comparison with the classical ANN, the network architecture was adjusted to approximate the number of trainable parameters in the optimized QC-REUP algorithm (approximately 360). Despite this adjustment, the ANN achieved significantly higher classification performance with a median F1 score of 0.75 (95% CI: 0.73-0.83). These findings suggest that while quantum classification circuits with data re-uploading can be configured to perform comparably to linear functions and can be trained on data with a larger number of features than other QML algorithms, classical non-linear functions such as ANNs still achieve better classification performance, even under ideal, noiseless quantum conditions. While it is thought that QC-REUP algorithms can universally approximate complex continuous functions, further research with higher dimensional datasets may clarify the limits of this capability (19,28).

A key objective of this study was to assess the capabilities of the QC-REUP algorithm as compared to QML and ML counterparts, using datasets with dimensionality and complexity comparable to those likely encountered in real-world healthcare applications. While QML algorithms, particularly QC- REUP, showed potential on lower-dimensional datasets, the results on higher-dimensional datasets suggest that QML is not yet ready to replace or surpass classical ML in this context. Classical algorithms, such as XGBoost, consistently performed better, indicating that laboratory medicine should continue to rely on classical ML approaches for practical applications and translational research. However, some literature has begun to explore hybrid QML-classical boosting algorithms with promising results (33). As quantum algorithm design and discovery continue to advance, it is possible that QML will improve on some of its current limitations. Therefore, it remains prudent for healthcare and laboratory medicine to continue evaluating this rapidly evolving technology as algorithm design and development advances (8).

This study had several limitations. Our first limitation related to assessing the impact of noise on QML classification performance. Noise refers to unintended disturbance of quantum states, such as superposition and entanglement, which can lead to degradation of information encoded in physical qubits and subsequent computational errors (8). While the simulators used in this study can simulate noise, runs with quantum noise models were excluded in our final analysis. This was due to computational resource constraints that underscore the practical limitations faced when simulating QML on classical hardware. A second limitation was that all QML algorithms were tested on quantum simulators rather than actual QPUs. Although QPUs are increasingly accessible through commercial cloud providers, conducting additional experiments using them was beyond this paper’s scope due to the additional optimization needed for QPU mapping and the financial constraints associated with limited resource access. Lastly, the PAA dataset employed in this study, while having higher dimensionality, may not fully represent the complexity of data encountered in other real-world healthcare, or laboratory medicine applications, thereby limiting the generalizability of the conclusions drawn from this study.

Future ML research in healthcare and laboratory medicine should include the evaluation of QML algorithms on both simulators and QPUs as they become more accessible and capable. Additionally, the investigation of emerging quantum algorithms, such as hybrid quantum-classical models, may provide insights for practical healthcare applications. The findings of this study highlight the potential of QML, which, despite being in the early stages of development, has shown comparable performance to classical ML algorithms in certain contexts. As quantum hardware and algorithm design advance, these results underscore the importance of continued evaluation of this technology. In the near term, QML may complement classical ML, particularly for tasks involving lower-dimensional datasets or hybrid frameworks. Over the long term, QML could become a significant addition to machine learning practices, but considerable technological and methodological advancements are likely required to fully realize its potential.

## Data Availability

All data produced are available online at: Edmund H Wilkes, Erin Emmett, Luisa Beltran, Gary M Woodward, Rachel S Carling, A Machine Learning Approach for the Automated Interpretation of Plasma Amino Acid Profiles, Clinical Chemistry, Volume 66, Issue 9, September 2020, Pages 1210-1218, https://doi.org/10.1093/clinchem/hvaa134

