## supplemental-material for "Quantum Machine Learning and Data Re-Uploading: Evaluation on Benchmark and Laboratory Medicine Datasets"

**Supplemental Figure 1:** Scatterplot of circle dataset by class.

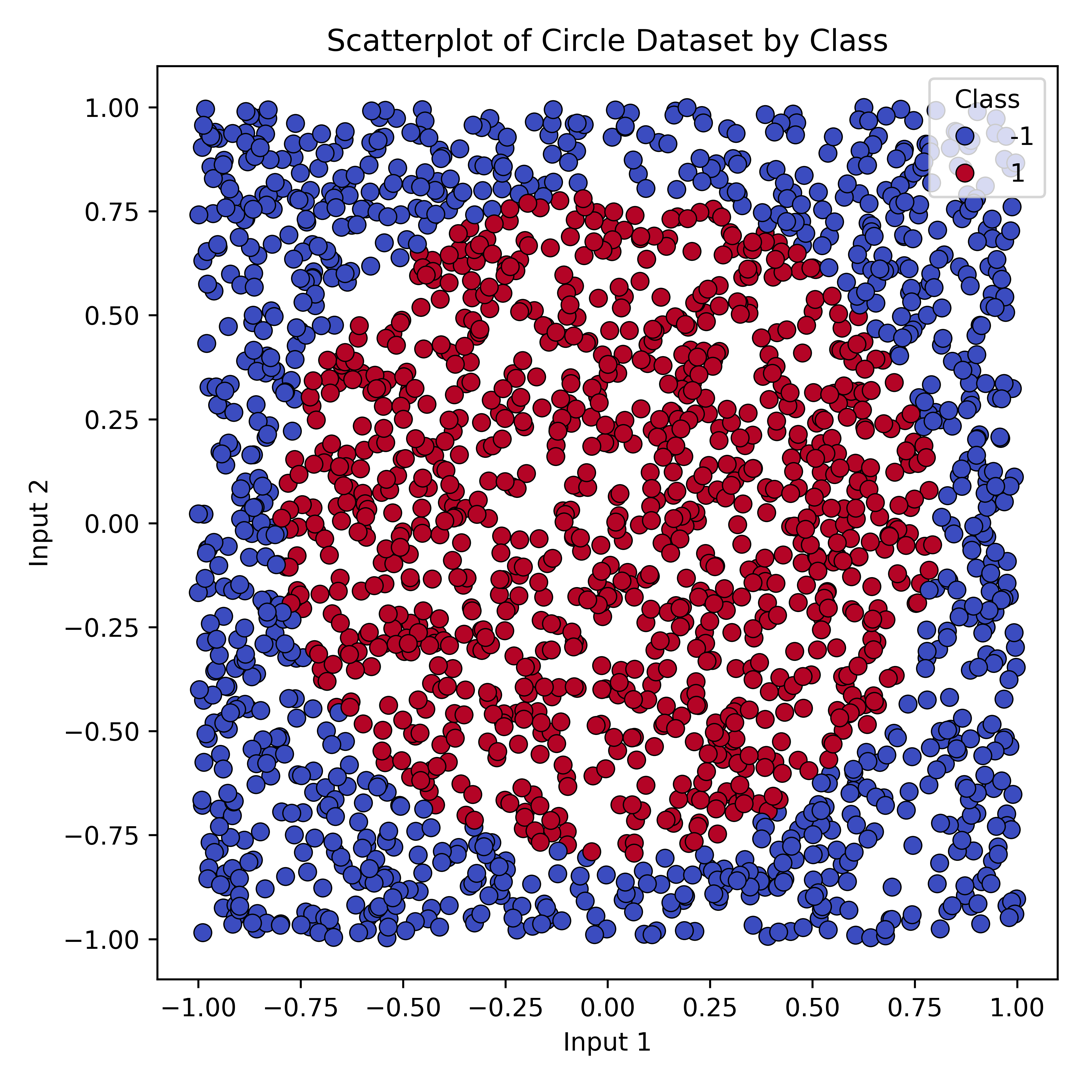

**Supplemental Table 1**: Mean (SD) accuracy for quantum re-upload on circle dataset with 3-fold cross validation.

| Number of Qubits | 1 | 2 | |
| --- | --- | --- | --- |
| Number of Layers |  | *Without Ent.* | *With Ent.* |
| *1* | 0.82 (0.00) | 0.78 (0.03) | 0.80 (0.03) |
| *2* | 0.85 (0.07) | 0.92 (0.01) | 0.93 (0.01 |
| *3* | 0.89 (0.01) | 0.89 (0.03) | 0.92 (0.00) |
| *4* | 0.84 (0.08) | 0.90 (0.01) | 0.93 (0.01) |
| *5* | 0.91 (0.01) | 0.84 (0.00) | 0.91 (0.01) |
| *6* | 0.90 (0.03) | 0.83 (0.03) | 0.90 (0.02) |
| *7* | 0.93 (0.00) | 0.87 (0.03) | 0.91 (0.02) |
| *8* | 0.92 (0.01) | 0.83 (0.01) | 0.94 (0.01) |

**Supplemental Figure 2**: Mean training loss and 95% confidence interval for the quantum re-upload algorithm on the plasma amino acid dataset.

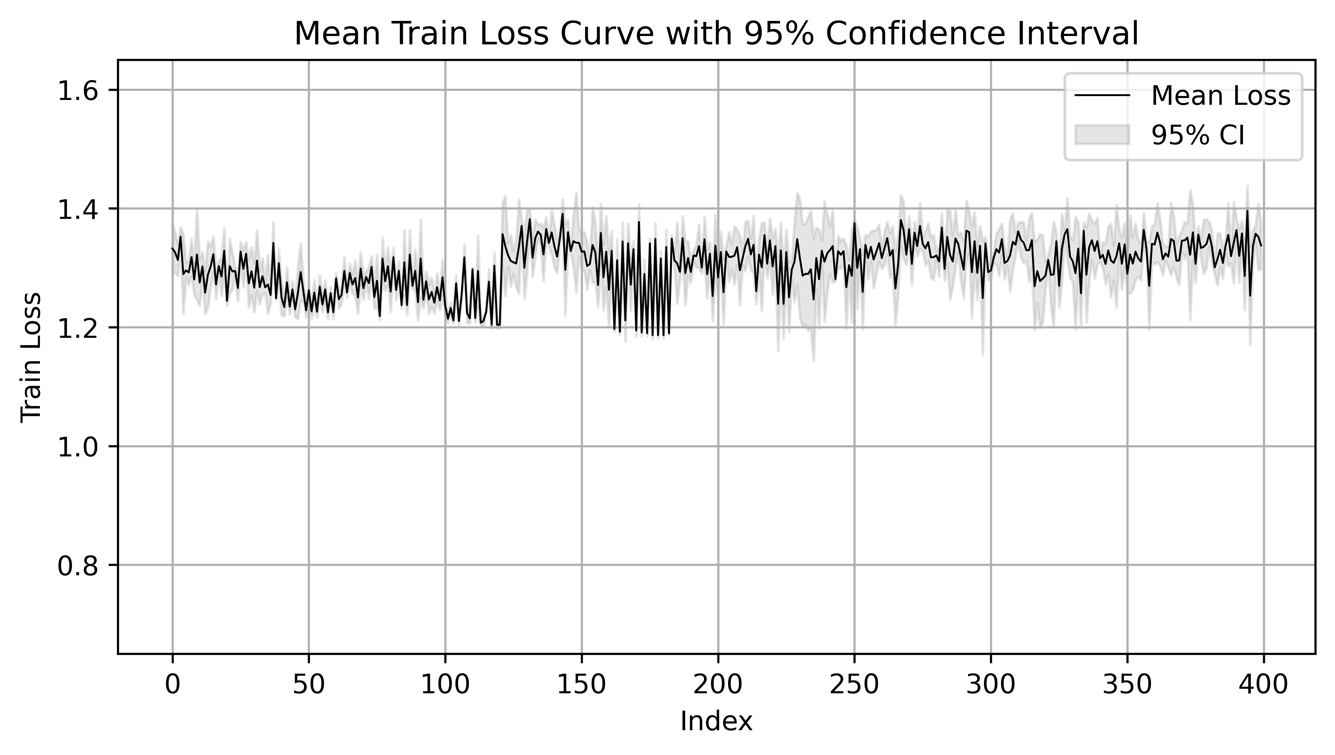

**Supplemental Figure 3**: Quantum circuit diagram for QC-REUP in Experiment 3

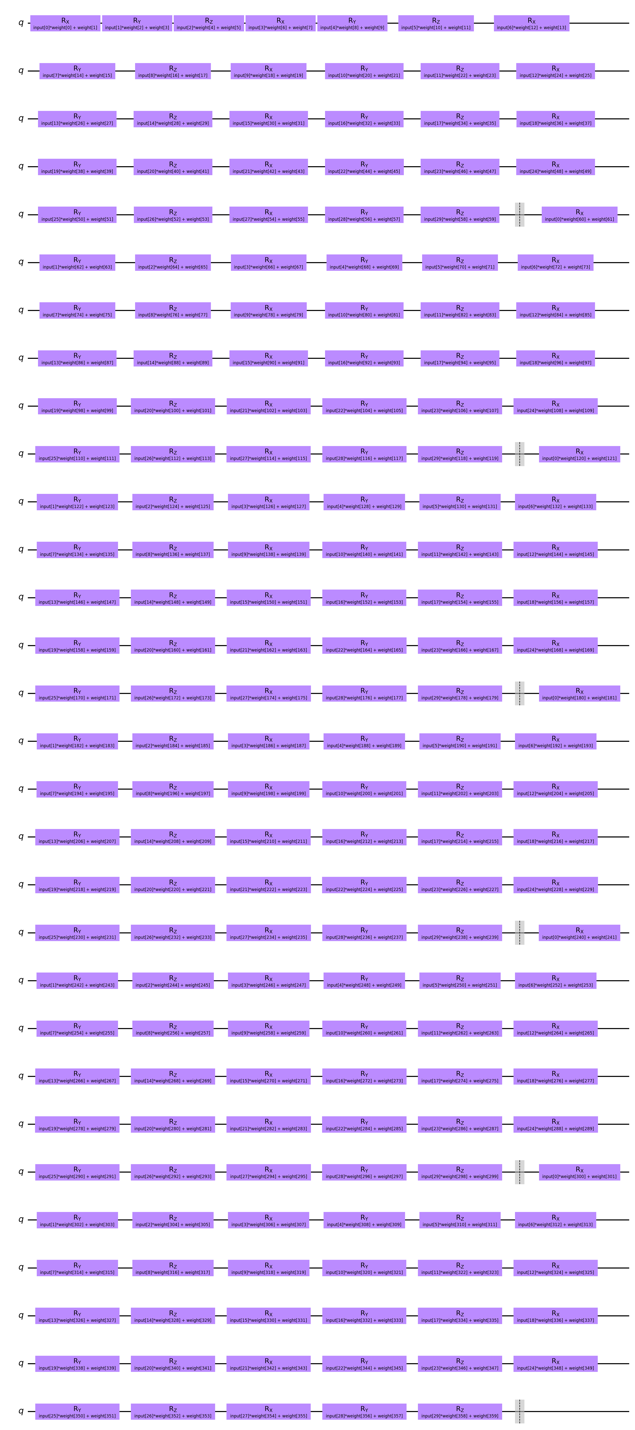

**Supplemental Table 2:** Importance of configuration parameters on F1 score for QC-REUP by random forest regression.

|  | importance |
| --- | --- |
| feature_scaler_min0max1 | 0.233726 |
| downsample_True | 0.17713 |
| q_num_qubit | 0.175506 |
| feature_scaler_no_scaler | 0.128975 |
| downsample_False | 0.081104 |
| q_num_layers | 0.0704 |
| feature_scaler_standard | 0.038733 |
| q_entanglement_one-qubit | 0.024548 |
| q_reup_method_asymmetrical | 0.018111 |
| q_entanglement_False | 0.017582 |
| q_reup_method_one-qubit | 0.012972 |
| q_reup_method_symmetrical | 0.012138 |
| q_entanglement_True | 0.009074 |
